# Vaccinating Australia: How long will it take?

**DOI:** 10.1101/2021.02.02.21250979

**Authors:** Mark Hanly, Tim Churches, Oisín Fitzgerald, C Raina McIntyre, Louisa Jorm

## Abstract

The Australian Government’s COVID-19 vaccine rollout strategy is scheduled to commence in late February 2021 and aims to vaccinate the Australian adult population by the end of October 2021. The task of vaccinating some 20 million people within this timeframe presents considerable logistical challenges. Key to meeting this target is the rate of vaccine delivery: the number of vaccine doses that can be administered per day. In the opening phase, high priority groups will receive the Pfizer/BioNTech vaccine through hospital hubs at an initial rate of 80,000 doses per week. However, pending regulatory approval, the currently announced plan appears to be to distribute the AstraZeneca vaccine to the bulk of the popluation through a combination of general practices and community pharmacies. Here, we run a series of projections to estimate how long it will take to vaccinate the Australian population under different assumptions about the rate of vaccine administration as well as the schedule for second doses and prevalence of vaccine hesitancy. Our analysis highlights the ambitious rate of vaccine administration that will be neccessary to meet the Australian Government completion target of October 2021. A rate of 200,000 doses per day would comfortably meet that target; 80,000 doses a day would see roll-out extended until mid-2022. Speed is of the essence when it comes to vaccine rollout: protecting the population quickly will minimise the risk of sporadic and costly lockdowns lockdowns and the potential for small, local clusters getting out of control and sparking new epidemic waves. The government should gather all its resources to maximise the daily vaccination rate, ideally aiming to ramp up administration to at least 200,000 doses per day as quickly as possible. Quickly achieving and maintaining this pace will likely require dedicated large-scale vaccination sites that are capable of delivering thousands of doses a week in addition to the enthusiastic participation of GP practices and community pharmacies around the country. Lessons on the neccessary logistical planning, including coordination of delivery, ultra-cold-chain storage and staffing, can potentially be learned from Israel, where between 7,000 and 20,000 vaccinations per million population have been delivered daily throughout January.

## 1 Introduction

The development and regulatory approval of multiple safe and efficacious COVID-19 vaccines in less than a year is a truly remarkable achievement. The logistical task of administering the vaccine rapidly and fairly to billions of people around the world will be no less of a challenge. National vaccination programs have commenced in many countries including Israel, the United States and the United Kingdom. In Australia, the federal government has entered into four agreements for the supply of COVID-19 vaccines (Table 1), with a view to starting distribution in late February 2021, and setting an ambitious target of completion of vaccination of the adult population in October 2021.^1^ This target allows just 35 weeks to administer two doses each to some 20 million adult Australians.

**Table 1:** Vaccine supply agreements entered into by the Australian government

| Name | Type | Doses<br>(millions) | Schedule | Status as at 1st February,<br>2021 |
| --- | --- | --- | --- | --- |
| Pfizer/BioNTech | mRNA vaccine | 10 | 2 doses, 21<br>days apart | Provisionally approved by<br>the TGA |
| University of<br>Oxford<br>AstraZeneca | Viral vector vaccine | 54 | 2 doses, 28<br>days apart | Phase 3 clinical trials |
| Novavax | Protein vaccine | 51 | 2 doses, 21<br>days apart | Phase 3 clinical trials |
| COVAX Facility | Assorted | 25 | Assorted | Nine candidate vaccines in<br>various clinical trial stages |

Adapted from https://www.health.gov.au/initiatives-and-programs/covid-19-vaccines/about-covid-19-vaccines/australias-vaccine-agreements

The national roll-out strategy divides the population into 16 groups, organised into five distinct phases (Table 2). Hospital hubs with access to −70^°^C ultra-cold-chain storage facilities will administer the Pfizer/BioNTech vaccine to the highest priority groups scheduled in Phase 1a, which includes border workers, frontline healthcare staff, and aged care staff and residents.^2^ Pending approval, the AstraZeneca vaccine would be administered to the bulk of the adult population through a network of general practitioners (GPs) and community pharmacies.

**Table 2:** Australia’s COVID-19 vaccine national roll-out strategy

| Phase | Description | Size |
| --- | --- | --- |
| 1a | Quarantine & border workers | 70,000 |
| 1a | Frontline health care workers | 100,000 |
| 1a | Aged care and disability care staff | 318,000 |
| 1a | Aged care and disability care residents | 190,000 |
| 1b | Elderly adults aged 80 years and over | 1,045,000 |
| 1b | Elderly adults aged 70-79 years | 1,858,000 |
| 1b | Other health care workers | 953,000 |
| 1b | Aboriginal and Torres Strait Islander people aged 55 years and over | 87,000 |
| 1b | Younger adults with an underlying medical condition | 2,000,000 |
| 1b | Critical and high risk workers | 196,000 |
| 2a | Adults aged 60-69 | 2,650,000 |
| 2a | Adults aged 50-59 | 3,080,000 |
| 2a | Aboriginal and Torres Strait Islander people aged 18-54 | 387,000 |
| 2a | Other critical and high risk workers | 453,000 |
| 2b | Balance of adult population | 6,643,000 |
| 3 | <18 if recommended | 5,670,000 |

Adapted from https://www.health.gov.au/sites/default/files/documents/2021/01/australia-s-covid-19-vaccine-national-roll-out-strategy.pdf

The Pfizer/BioNTech and AstraZeneca vaccines both require two doses—a primer and a booster—which need to be delivered within a specified time frame after the initial shot. This complicates roll-out as the resources of specialised vaccine administration facilities, and nominated general practices and pharmacies must be divided between unprotected individuals waiting for their first injection and those who have already been afforded some protection and who are returning for their booster administration. Furthermore, the duration of protection afforded by these vaccines is not yet well characterised, and re-vaccination of some or all of the population may be required. We have not included re-vaccination into the any of the scenarios examined in this paper, at this stage.

Another unknown is the question of vaccine hesitancy, which refers to delay in acceptance or complete refusal of vaccination, despite a suitable vaccine being available and accessible.^3^ Clearly, high levels of vaccine hesitancy would have the potential to undermine efforts to establish adequate protection of the whole population through herd immunity. An online survey of over 3,000 Australian adults undertaken in August 2020 asked respondents if they would agree to vaccination for COVID-19 if a safe and effective vaccine were available. The population-weighted responses were 5.5% *definitely not*, 7.2% *probably not*, 28.7% *probably yes* and 58.5% *definitely yes*.^4^

When it comes to vaccine roll-out, speed is of the essence. Statistical modelling has illustrated that epidemic duration, cases and deaths are minimised dramatically as the number of available daily vaccinations increases. In one modelling scenario, for example, increasing the daily capacity by 25% from 75,000 to 100,000 resulted in a 60% reduction in total cases and deaths.^5^ The Australian Prime Minister has cited a schedule starting at 80,000 vaccinations per week and scaling up from there.^6^ It is unclear what the peak daily vaccination target is, but a simple “back of the envelope” calculation suggests that in order to vaccinate some 20 million adult Australians twice in the eight months from the start of March 2021 to the end of October 2021 would take in the order of 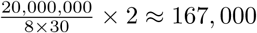 doses per day.

The aim of this analysis is to estimate how long it might take to administer the announced two-dose COVID-19 vaccine schedule to the Australian population. We consider a variety of scenarios based on the daily vaccine administration capacities, the timeframe between the first and second doses and the scale of vaccine hesitancy in the population. We conclude by comparing daily vaccine administration rates from countries where vaccination programs are already underway.

## 2 Methods

### 2.1 Population and priority groups

Our analysis is based on the 16 priority groups and five phases proposed by the Australian government (see Table 2). The assumed population size is 25.7 million people, including 5.67 million children and adolescents under the age of 18. We also assume that equal priority will be given to all groups within the same phase.

### 2.2 Vaccine roll out projections

Roll out projections are based on three parameters:

1. The daily vaccination capacity.
2. The valid window (in days) to receive the second dose after the first has been administered.
3. Vaccine hesitancy.

### 2.3 Projection scenarios

Projection scenarios are based on a 2^*k*^ factorial design defined by three factors with two levels each. The scenarios are summarised in Table 3 and the three factors and levels are detailed below:

**Table 3:**
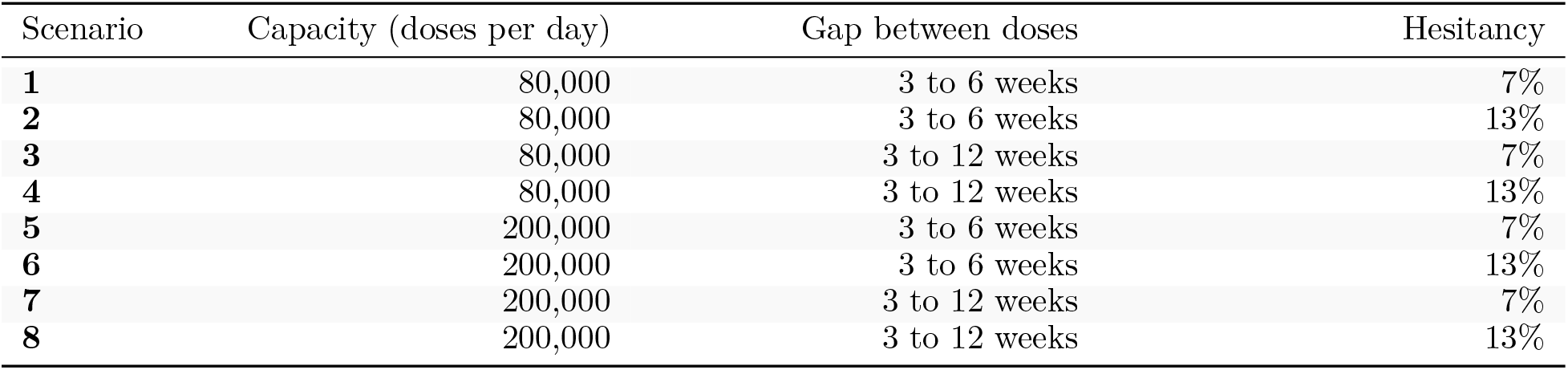
Projection scenarios

**Table 4:**
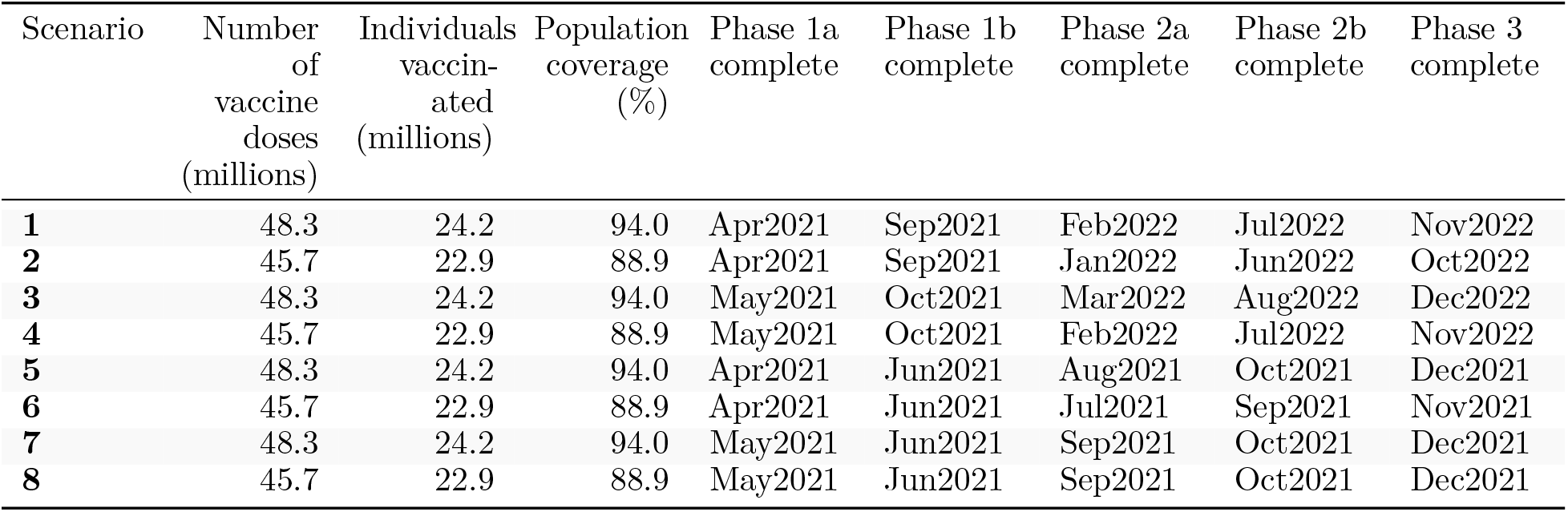
Summary of vaccine roll-out projections for different scenarios

- Daily vaccination capacity (80,000 versus 200,000)
- Timing between first and second dose (3-6 weeks versus 3-12 weeks)
- Vaccine hesitancy (7% versus 13%).

The projections assume that those who are hesitant will never receive the vaccine. The specified hesitancy rates are based on the survey data reported by Edwards *et al*.,^4^ and are only applied to general population groups. Border staff, healthcare and aged care workers, aged care residents and adults with a medical condition were assumed to have negligible vaccine hesitancy.

### 2.4 Vaccine allocation

We allocated the daily available vaccination doses according to the following algorithm:

1. Calculate the number of second doses due, based on the specified permissible range for the second dose. As an example, if the second dose is specified to be administered at between three and six weeks, then the booster doses for people vaccinated on day 1 would be evenly distributed across the three weeks between day 22 and day 42).
2. Assign the remaining doses from the daily limit to those awaiting their first dose.
3. Identify the highest priority phase that hasn’t yet received all first doses.
4. Divide the available first doses between the subgroups in the highest priority phase, proportional to the number of unvaccinated individuals remaining in each subgroup.
5. Stop when all population members, minus those who are hesitant, have been vaccinated twice.

### 2.5 Software and code

The analysis was performed using R version 4.0.3^7^ and associated packages^8^. The complete source code to reproduce this analysis can be accessed at https://github.com/CBDRH/vaccinatingAustralia.

## 3 Results

Results from the eight scenarios are presented in Table 3. Scenarios that assumed a vaccine hesitancy of 7% among the general population resulted in 48.3 million vaccine doses administered to 24.2 million people, corresponding to a population coverage of 94.0%. Scenarios that assumed the vaccine higher hesitancy rate of 13% resulted in 45.7 million vaccine doses administered to 22.9 million people for a coverage of 88.9%.

Under an optimistic Scenario of 200,000 daily doses with the 7% hesitancy rate (Scenario 5), assuming a start date of 1 March 2021, Phase 1a would be fully vaccinated (i.e. primer and booster doses administered) as early as 13 April 2021—in just six elapsed weeks. The entire adult population would be fully vaccinated by about 25 October 2021—in line with government targets—and a further eight weeks would see the entire population include those under 18 vaccinated. Under this optimistic scenario, the vulnerable groups in Phase 1b, including adults aged 70 and above and those with underlying medical conditions, would be fully vaccinated before the onset of the Southern Hemisphere winter. Under this scenario, we would reach 50% population coverage in late July 2021 and 75% population around the start of October 2021 (Figure 1A).

**Figure 1:**
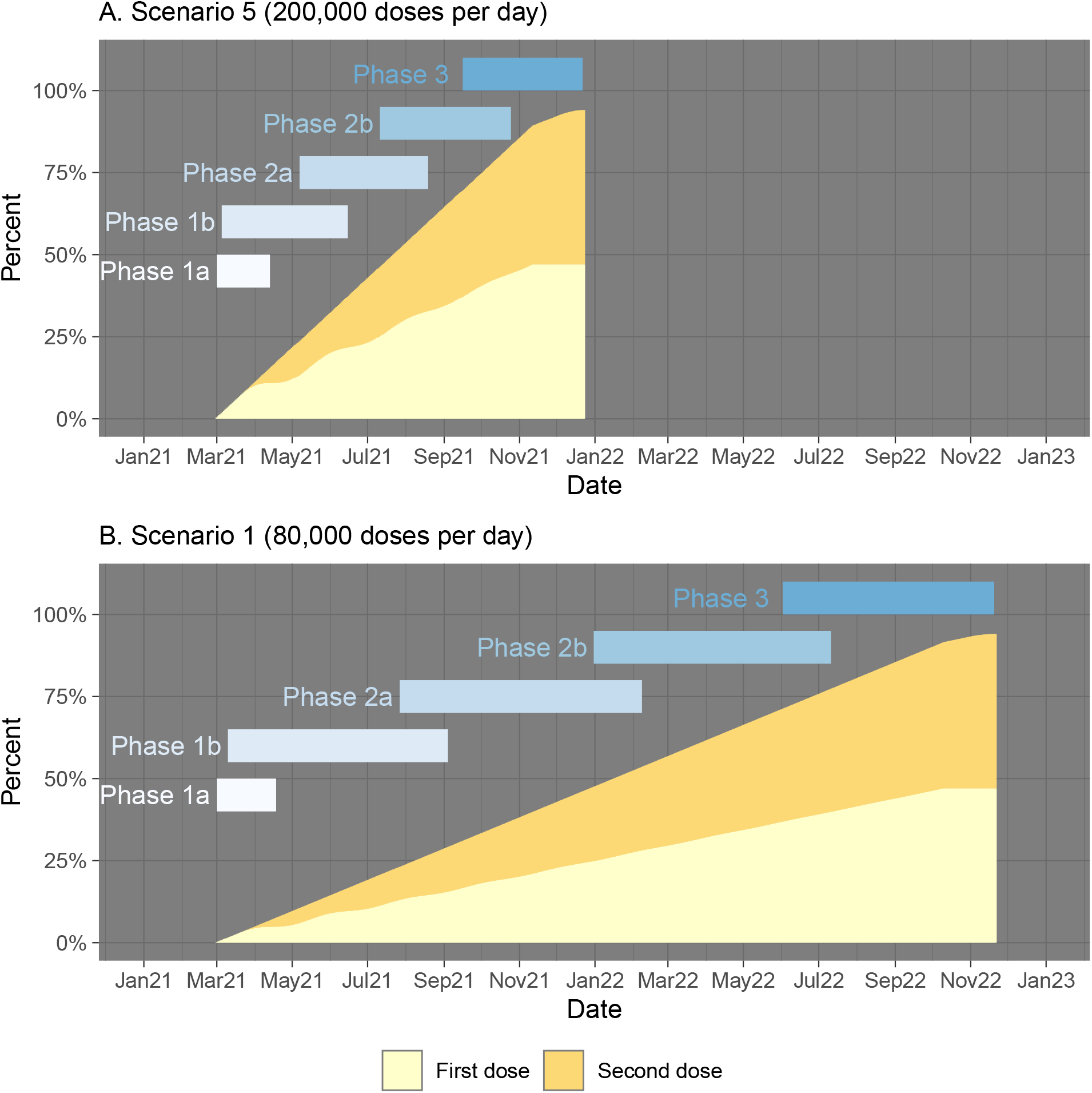
Cumulative vaccine doses over time.

Under the less optimistic scenarios of 80,000 doses administered daily, it would take until some time between June and August 2022 to vaccinate the adult population (Table 3). Under Scenario 1, we would reach 50% population coverage in January 2022 and 75% population coverage in July 2022 (Figure 1B).

## 4 Discussion

To meet the target of vaccinating all adult Australians by the end of October 2021 there will need to be, on average, in the order of 200,000 doses delivered daily (including weekends and holidays)–a truly furious pace.

To test the feasibility of such a vaccination administration rate, it is illuminating to consider the COVID-19 vaccination rates currently being reported by other countries which have already begun to roll out their vaccination programs (Figure 2). The outlier is Israel, where between 7,000 and 20,000 vaccinations per million population have been delivered daily throughout January. Several factors may have contributed to this success, including a young, largely urbanised population and a strong public health infrastructure. Perhaps most important has been strong logistical planning, including coordination of delivery, ultra-cold-chain storage and staffing.^9^ Other countries have been less successful in their roll-out to date, including the United States (∼4,000 per million pop max), the United Kingdom (∼5,000 per million pop max) and the European Union (∼1,100 per million pop max).

**Figure 2:**
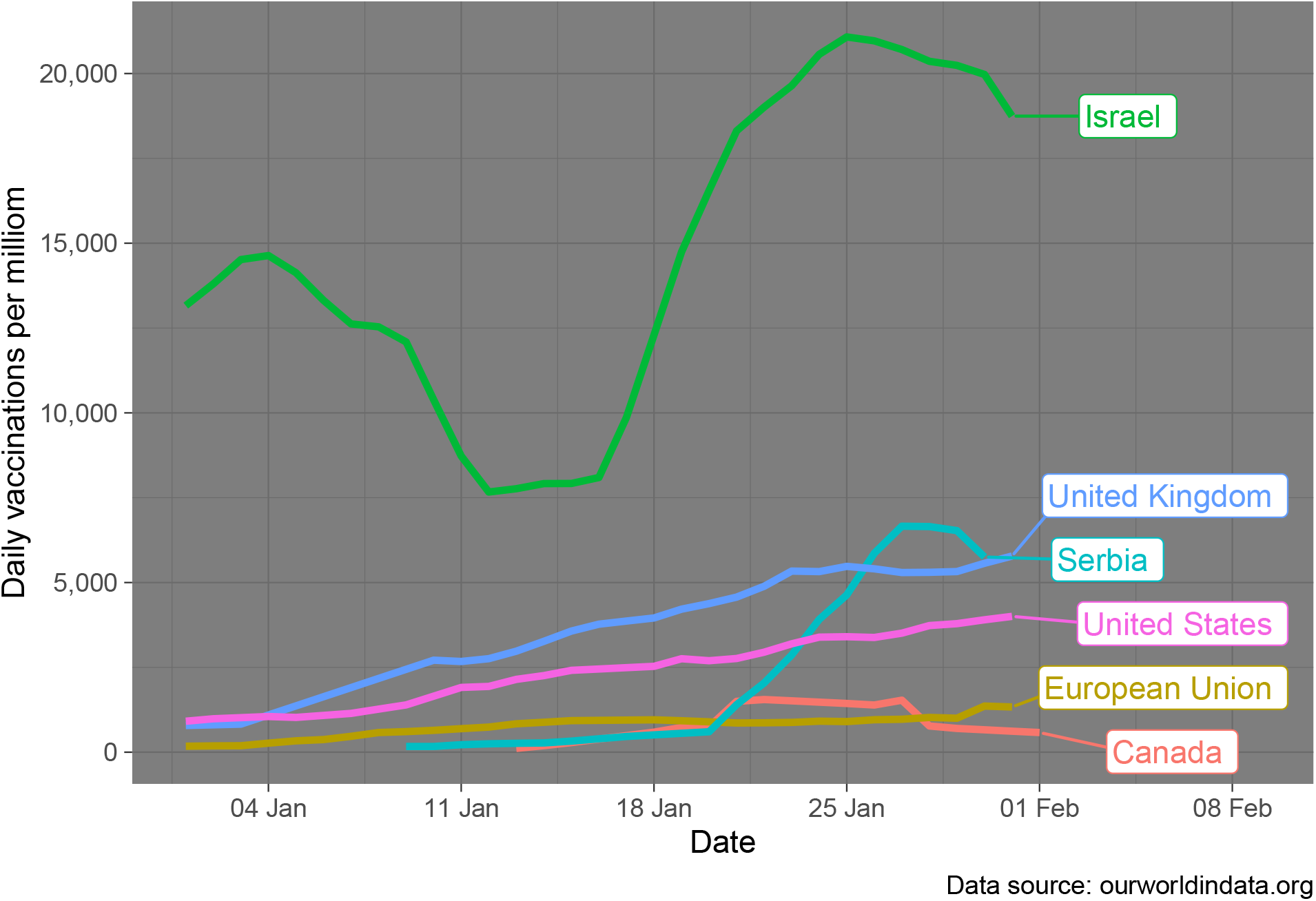
Daily COVID-19 vaccines administered per one million population.

With a population of 25.7 million in Australia, the figure of 200,000 doses per day from our projections corresponds to approximately 7,800 daily doses per million population. Thus, it does seem possible to vaccinate the Australian population in just eight months, but administration rates considerably better than those currently being achieved in most countries will be needed. Clearly Australia needs to start as soon as possible, rapidly ramp-up to the required vaccination velocity and then to maintain that relentless pace.

Apart from Phase 1a, currently announced delivery plans involve recruiting, authorising and training 1,000 general practices and an unknown number of pharmacies to administer vaccine doses, as well as capture and record the required documentation for each vaccinee.^10^ If authorised general practices alone are relied upon to vaccinate the bulk of the population from Phase 1b onwards, then on average each practice would need to administer 200 doses per day, seven days per week, for about six months. This seems infeasible for all but the largest group practices. There are some 5,800 community pharmacies across Australia.^11^ If we assume that half of these are willing and able to participate in the vaccination programme, and each delivers an average of 35 doses per day (including weekends and holidays), then this would represent half of the daily administration capacity of 200,000 doses per day to meet the whole of community vaccination target of the end of October 2021. However, that still means that the 1,000 authorised general practices would need to administer an average of 100 doses per day. This would still seem to be a stretch, given that those practices would still have their normal workloads to contend with. It seems clear that to deliver at the scale needed will require dedicated large-scale vaccination sites that are capable of delivering several thousands of doses a week, in addition to the enthusiastic participation of GP practices and community pharmacies around the country.

Of course, there are also major logistical challenges in ramping up such a vaccination capacity. Adapting existing supply chain and vaccine information systems to meet the demand is likely to take several weeks at the most optimistic, and adequately training several clinical and clerical staff from each of the authorised vaccination general practices and pharmacies to safely screen patients, deliver the vaccines and record the results is likely to take much longer than just one month. Thus our assumption of commencement of vaccine roll-out at full velocityby March 2021 seems particularly optimistic, and therefore the estimates reported in the tables above are likely to be similarly optimistic.

## 5 Contributions

MH, TC and LJ conceived of the study; MH, OF and TC wrote the R code; MH drafted the manuscript; all authors reviewed and edited the manuscript.

## Data Availability

Data generated through our projections are hosted in the GitHub repo for this project.
Data on the daily vaccinations per million population were accessed through ourworldindata.org

https://ourworldindata.org/covid-vaccinations#how-many-covid-19-vaccine-doses-are-administered-daily

## 6 Acknowledgements

This research was supported by the generous assistance of Ian Sharp, philanthropic supporter of UNSW research, and by a research seed grant provided by the Sydney Partnership for Health, Education, Research and Enterprise (SPHERE) Infectious diseases, Immunity and Inflammation (Triple-I) Clinical Academic Group.

## Notes

### Competing Interest Statement

Mark Hanly receives funding from the National Health and Medical Research Council (NHMRC) and the Sydney Partnership for Health, Education, Research and Enterprise (SPHERE).
Timothy Churches receives funding from the Sydney Partnership for Health, Education, Research and Enterprise (SPHERE), Australian Research Data Commons (ARDC), UNSW Sydney and the Ingham Institute for Applied Medical Research.
Oisin Fitzgerald receives funding from the Sydney Partnership for Health, Education, Research and Enterprise (SPHERE).
C Raina MacIntyre receives funding from Medical Research Futures Fund and NHMRC. She has consulted for Astra Zeneca and Janssen on COVID-19 vaccines and been on an advisory board for Seqirus on COVID-19 vaccines. In the past she has been on advisory boards and had support from vaccine manufacturers Sanofi, Seqirus, Merck, Pfizer, GSK, Bavarian Nordic and Emergent Biosolutions for other vaccines.
Louisa Jorm receives funding from the Medical Research Futures Fund, Australian Research Data Commons, NHMRC, UNSW Sydney and the Sydney Partnership for Health, Education, Research and Enterprise (SPHERE).

### Author Declarations

N/A - only published data was used in this mathematical modelling study.

### Summary of Updates

Authorship list updated.

## References

1. Health AGD of. Doorstop interview on 31 january 2021. Australian Government Department of Health. Published online January 2021. https://www.health.gov.au/ministers/the-hon-greg-hunt-mp/media/doorstop-interview-on-31-january-2021

2. Australia’s covid-19 vaccine national roll-out strategy. Published online December 2020. https://www.health.gov.au/sites/default/files/documents/2021/01/australia-s-covid-19-vaccine-national-roll-out-strategy.pdf

3. MacDonald NE, others. Vaccine hesitancy: Definition, scope and determinants. Vaccine. 2015;33(34):4161-4164.

4. Edwards B, Biddle N, Gray M, Sollis K. COVID-19 vaccine hesitancy and resistance: Correlates in a nationally representative longitudinal survey of the australian population. medRxiv. Published online 2020.

5. MacIntyre CR, Costantino V, Trent MJ. Modelling of covid-19 vaccination strategies and herd immunity, in scenarios of limited and full vaccine supply in nsw, australia. medRxiv. Published online 2020.

6. Press conference - australian parliament house. Published online January 2021. https://www.pm.gov.au/media/press-conference-australian-parliament-house-12

7. R Core Team. R: A Language and Environment for Statistical Computing. R Foundation for Statistical Computing; 2019. https://www.R-project.org/

8. Wickham H, Averick M, Bryan J, et al. Welcome to the tidyverse. Journal of Open Source Software. 2019;4(43):1686. doi:10.21105/joss.01686

9. McKee M, Rajan S. What can we learn from israel’s rapid roll out of covid 19 vaccination? Israel Journal of Health Policy Research. 2021;10(1):1–4.

10. Health AGD of. GPs’ key role in covid-19 vaccination rollout. Australian Government Department of Health. Published online January 2021. https://www.health.gov.au/ministers/the-hon-greg-hunt-mp/media/gps-key-role-in-covid-19-vaccination-rollout

11. Vital facts on community pharmacy. Published online December 2020. https://www.guild.org.au/data/assets/pdf_file/0028/91909/PGA_December_2020_infographic.pdf

